# Estimation of Myocardial and Blood Gadolinium Concentrations from T1 Mapping via Pharmacokinetic Modeling: Influence of Elastic Deformation Registration

**DOI:** 10.1101/2025.10.02.25337032

**Authors:** Yasutoshi Ohta, Masaru Shiotani, Yoshiaki Morita, Tatsuya Nishii, Hiroki Horinouchi, Akiyuki Kotoku, Midori Fukuyama, Emi Tateishi, Tetsuya Fukuda

**Author notes:** **Corresponding author**: Yasutoshi Ohta M. D, Ph. D, Department of Radiology, National Cerebral and Cardiovascular Center, Suita City, Osaka 564-8565, Japan.

## Abstract

Accurate quantification of myocardial gadolinium-based contrast medium (CM) concentration is important for longitudinal cardiac MRI assessment. We developed a T1 mapping-based pharmacokinetic (PK) approach incorporating elastic deformation registration (ER) to improve fitting accuracy. Forty-nine participants (median age, 62 years) underwent native and dynamic T1 mapping at 2, 5, 9, and 15 minutes after CM administration. CM concentration maps for myocardium and blood pool were generated at each time point, with and without ER, and fitted to a PK model. Model performance was evaluated using the coefficient of determination (R²), Akaike Information Criterion (AIC), and Bayesian Information Criterion (BIC). Agreement between measured myocardial CM concentrations and model estimates was assessed, and the mean difference between them was calculated. T1-derived CM concentrations showed excellent agreement with phantom reference values (R^2^ = 0.999). ER improved model fit compared with no registration, with smaller absolute mean differences at all time points: 0.026 vs. 0.029 mM/l (2 min), –0.021 vs. –0.025 (5 min), –0.022 vs. –0.023 (9 min), and 0.018 vs. 0.021 mM/l (15 min). This ER-based dynamic T1 mapping approach improved the accuracy of PK modeling for myocardial CM concentration, supporting its potential utility for refined myocardial tissue characterization.

## Introduction

Late myocardial enhancement (LGE) is an established technique for detecting myocardial fibrosis and offers critical insights into myocardial structure ^1,2^. However, LGE captures only a snapshot in time, and the dynamic nature of contrast medium (CM) distribution within the myocardium may lead to missing subtle temporal variations. The conventional inversion recovery method, which nulls the signal from normal myocardium to emphasize lesions, often fails to detect nuanced changes, particularly over time. This limitation is particularly pronounced in conditions such as amyloidosis or dilated cardiomyopathy, where lesions are diffusely heterogeneous and pose challenges for inversion recovery (IR)-based assessments.

In recent years, advancements in myocardial imaging have introduced quantitative indices, such as T1 and T2 mapping and extracellular volume fraction (ECV) calculations, enhancing our ability to detect and quantify diffuse myocardial pathology. These advancements include the use of Modified Look-Locker Inversion recovery (MOLLI) sequences and motion correction techniques to improve the T1 map accuracy ^3,4^. Furthermore, developments in alignment methods for ECV calculation have significantly enhanced the quality of ECV maps, with techniques like elastic deformation registration (ER) providing better alignment ^5–7^, Additionally, the T1 values measured before and after contrast administration are now utilized to ascertain gadolinium CM concentrations^8^.

Gadolinium CM concentrations are known to decline biexponentially in the plasma following intravenous injection ^9^. Accurate in vivo measurement of the CM concentration in tissues with autonomous motion, such as the heartbeat of the myocardium, has been challenging because of technical limitations in aligning pre- and post-contrast images. We propose that aligning all T1 maps, both native and post-contrast, over time may offer a novel approach for myocardial tissue assessment. This method allows the calculation of CM concentration over time and enables a detailed evaluation of contrast kinetics at the voxel level. This study aimed to develop and explore the feasibility of myocardial pharmacokinetic analysis using ER techniques. As a potential clinical application, this method offers novel approaches for myocardial assessment, such as introducing a time-based axis for evaluating temporal changes and differentiating normal from abnormal myocardium based on CM concentration. Additionally, it allows for standardization of the post-contrast T1 mapping acquisition time across subjects and institutions when calculating extracellular volume (ECV), potentially improving inter-patient and inter-institutional comparability.

## Methods

### Phantom study

The contrast medium concentration phantom (Fig. 1a) was used to confirm the linearity and correlation between the diluted CM concentration and the CM concentrations calculated from T1 values. The CM (gadobutrol; Gadovist®, Bayer-Yakuhin, Osaka, Japan) was diluted in normal saline to the following concentrations (0.1, 0.2, 0.4, 0.5, 0.6, 1.0, 2.0, and 2.5 mM/l) and stored in 15 mL vials. The container containing the vial is filled with normal saline. The phantom was imaged using a 3T MRI system (MAGNETOM Vida; Siemens Healthcare, Erlangen, Germany) and a 30-channel body coil. T1 mapping was performed using a motion-corrected Look-Locker inversion recovery (MOLLI) sequence with 5(3)3 and 4(1)3(1)2 acquisition schemes, for nativeT1map and post-contrast T1map ^10,11^, respectively. The imaging parameters used were: TR/TE/flip angle, 3.8 msec/1.2 msec/35; slice thickness, 8 mm; voxel size, 1.4 ×1.4 ×8.0 mm^3^, NEX 1, FOV 360 x 306, matrix 256 x 144, parallel processing factor 2, initial effective inversion time 100msec with an increment of 80msec. ECG gating was used with a simulated ECG at 60 bpm from the system function. After imaging the vials at the center level of the phantom, T1 maps were automatically generated by the system. Circular regions of interest (19mm^2^) were placed at the center of the vials on images (Fig. 1b), and the T1 values for each concentration were measured. The native T1 value of normal saline was measured using native T1 MOLLI scheme (5(3)3), and the post-contrast MOLLI scheme (4(1)3(1)2) was used for the T1 value measurement of contrast medium-contained vials. The contrast medium concentration was then calculated using equation (Eq.1).

**Fig. 1.**
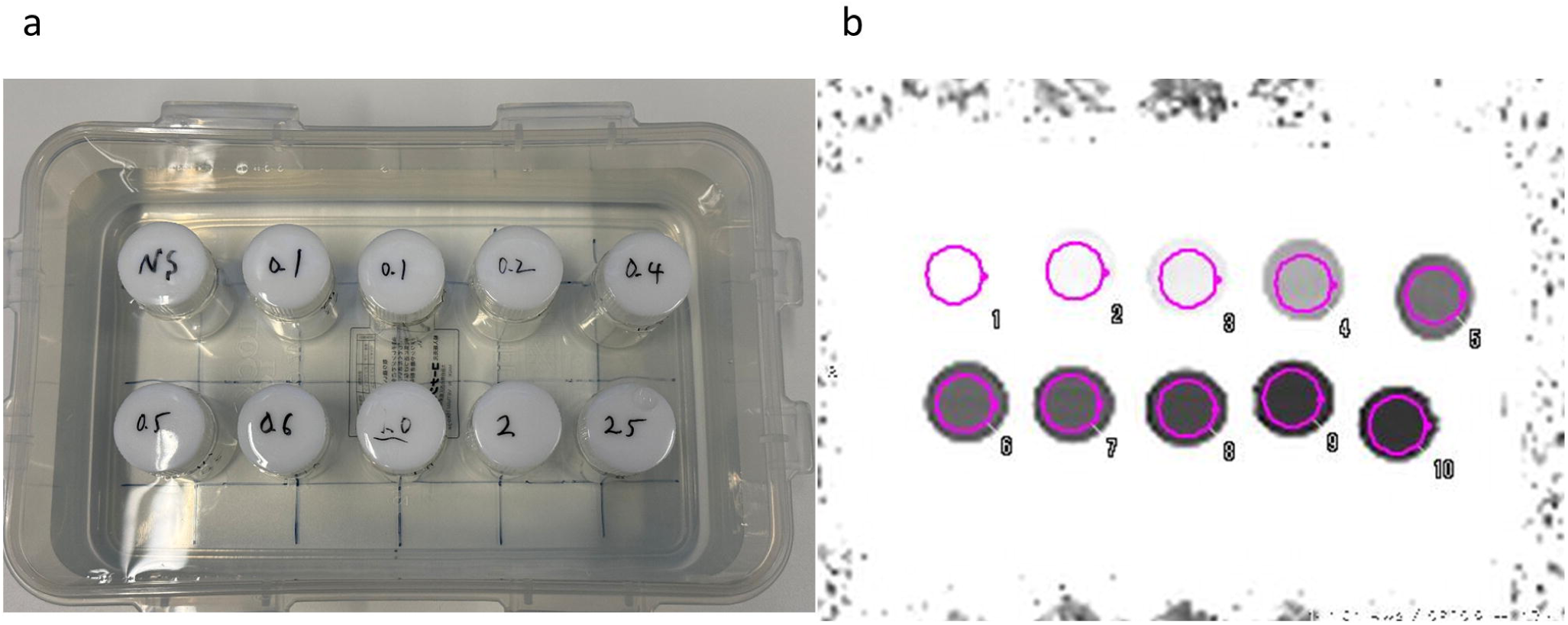
Photograph of the phantom (a) and T1 map (b) of diluted contrast medium. Diluted contrast medium is arranged in a container and the container is filled with normal saline. The region of interest (red circles) on the T1 map corresponds to the order of the vials in the photo.

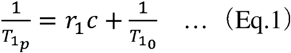

where 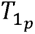 indicates the post-contrast T1 value of each time point, r_1_ indicates the relaxivity value of gadobutrol at 3.0 Tesla, c indicates concentration of contrast medium (mM/L), 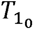 indicates native T1 value. The r_1_ value for gadobutrol varies slightly between reports^12^, and the r_1_ value of 5.0 given in the product monograph or report by the American College of Radiology was used ^13,14^.

### Study participants for in vivo evaluation

This single-center prospective study was approved by our institutional ethics committee (approval no. R20086). All methods were performed in accordance with the relevant guidelines and regulations. This study included subjects with known or suspected ischemic or non-ischemic cardiomyopathy who were referred to our radiology department between November 2022 and April 2023 for further myocardial characterization using cardiac MRI with contrast enhancement. Exclusion criteria included acute coronary syndrome, pregnancy, an estimated glomerular filtration rate < 30 mL/min/1.73 m^2^, claustrophobia, known allergy to gadolinium-based CM, and contraindication to MRI. No subjects met the exclusion criteria, and a total of 49 participants were included in the study. Written informed consent was obtained from all the participants before undergoing cardiac MRI.

### Cardiac MRI protocol

Cardiovascular magnetic resonance examinations were performed at our institution using a 3T MRI system (MAGNETOM Vida; Siemens Healthcare, Erlangen, Germany) with a 30-channel body coil. MR images were acquired as follows: cine imaging in all cardiac axis views, native T1 mapping, rest perfusion, dynamic T1 mapping (2, 5, 9, and 15 min after CM administration), and inversion recovery LGE (IR-LGE) true-FISP 10 min after contrast injection (Fig. 2a). Native and postcontrast T1 mappings were performed using a Look-Locker inversion recovery (MOLLI) sequence with 5(3)3 and 4(1)3(1)2 acquisition schemes, respectively. The acquisition parameters were as follows: TR/TE/flip angle, 3.8 msec/1.2 msec/35; slice thickness, 5 mm; voxel size, 1.4 × 1.4 × 5.0 mm^3^, NEX 1, FOV 360 × 306, matrix 256 × 144, parallel imaging factor 2, and an initial effective inversion time 100 msec with an increment of 80 msec ^15^. Three short-axis images (basal, mid, and apical) were obtained for each acquisition time. T1 maps were generated using in-line pixel-wise curve fitting with a three-parameter signal model for MOLLI inversion recovery with motion correction and an ER algorithm ^3,4^. To make our imaging protocol compatible with existing perfusion protocols for other research purposes, the CM dose was administered in two parts. A 0.05 mmol/kg of CM was injected for each perfusion imaging. The injection rate was 5 mL/sec with a 10-mL saline flush. The interval between perfusion scans was approximately 100 seconds, set based on the perfusion imaging time and the operation time. For LGE, the Look-Locker approach was used to determine the inversion time for nulling normal myocardial signal.

**Fig. 2.**
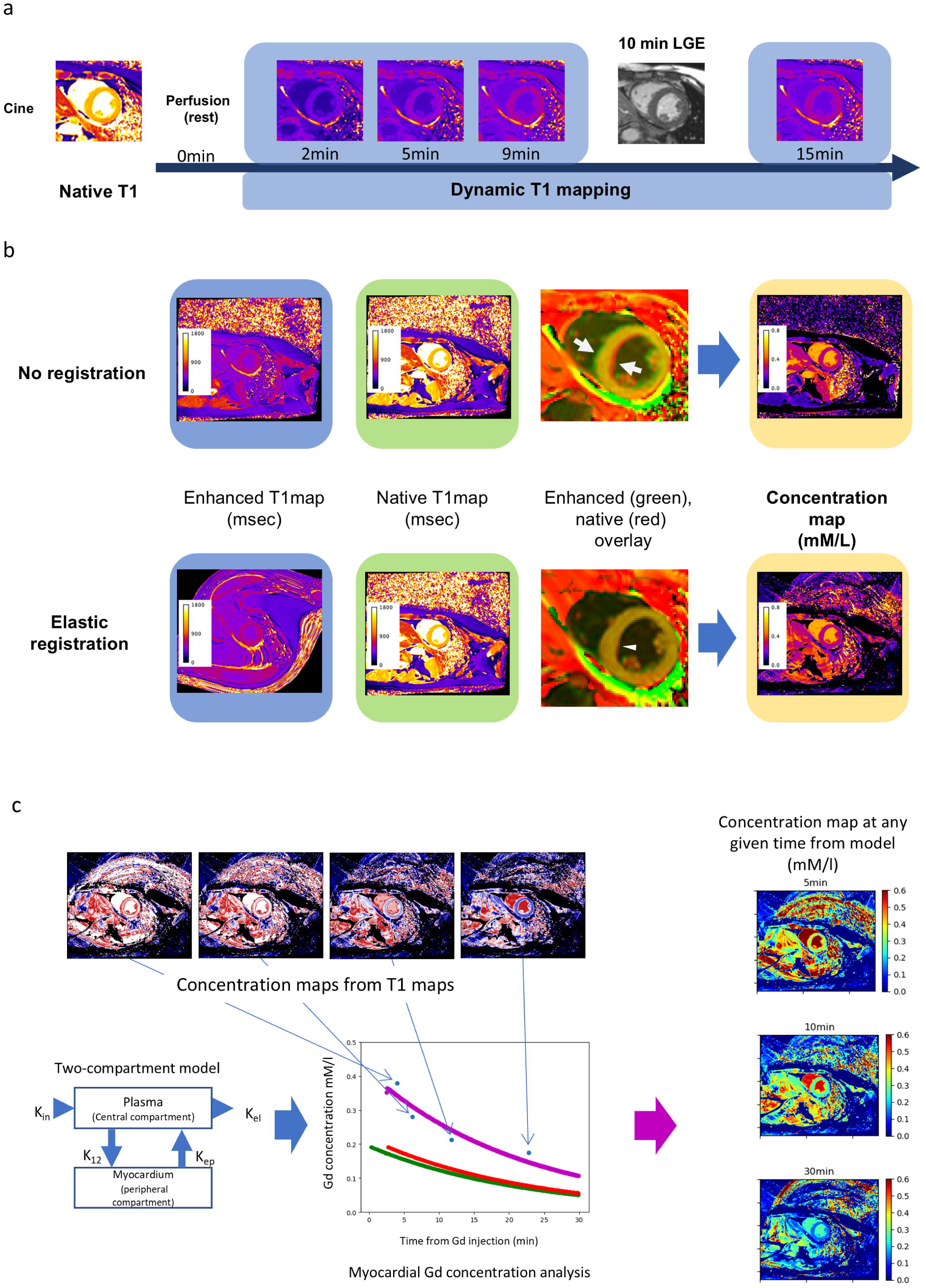
**a** Dynamic T1 mapping protocol. After cine images acquisition, native T1 maps were imaged. Following rest perfusion imaging, post-contrast T1 maps were obtained at 2, 5, and 9 min on the same slices as the native T1 map. LGE imaging was performed using the inversion recovery method at 10 min. T1 maps were acquired on the same slices as the native T1 map at 15 min. **b** At each time point after contrast administration, the concentration map was calculated from the post-contrast and native T1 maps. In the upper panel, the contrast medium concentration map was created without registration between the post-contrast and native T1 maps. Along the border of the left ventricular septum, an area of poor alignment (arrow) between the post-contrast map (green) and the native T1 map (red) is observed. In the lower panel, the concentration map was created after aligning the pre-contrast T1 map to the post-contrast T1 map using elastic registration in the same participant. In the overlay image, the myocardial boundaries match in both images, the entire myocardium is observed in orange (arrowhead), and no color blurring due to poor alignment of the myocardial boundaries is observed, as in the image without registration. **c** The process involves creating a predicted concentration map from the PK model. At each time point, curve fitting is performed for each pixel using a PK model based on a 2-compartment model applied to the concentration (blue dots). The post-administration time is accurately plotted using DICOM tags. Predicted concentration curves (first administration: green, second administration: red, superpositioned concentration curves: purple) are created for each pixel based on the parameters, resulting in the generation of a myocardial contrast medium concentration map at any given time (right column). *LGE* late gadolinium enhancement, *K_in_* rate constant equal to the infusion rate; *K_12_* rate constant for transfer of contrast medium from the central compartment; *K_el_* elimination constant of the contrast medium from the central compartment; *K_ep_* exchange rate constant from the extravascular extracellular space to plasma; *DICOM* digital imaging and communication in medicine.

### Image processing

The ER dataset was generated to minimize variations in cardiac chamber size caused by slight changes in cardiac phase at each imaging time point, as well as differences in diaphragm position due to breath-holding. This process ensured consistent myocardial alignment across all T1 maps. The registration was performed using the feature/contour-based registration function for ECV map creation on the workstation (cvi42, version 5.13.5, Circle Cardiovascular Imaging, Calgary, Canada), with the native T1 map used as the reference image. First, the left ventricular myocardial contour was automatically detected for both native and post-contrast T1 maps, and corrections were performed manually if there were apparent contour trace deviations. Next, ER was performed on the post-contrast T1 map images for each slice by applying the image registration function of the workstation. This was repeated for each post-contrast T1 map to align the images in all slices. Image registration using the workstation was not performed for the non-registration datasets.

The CM concentration maps for the ER and non-registration datasets (Fig. 2b) were generated using the voxel-by-voxel approach at all time points, applying Eq.1. An r_1_ value of 5.0 was utilized, as in the phantom study ^13,14^.

### Pharmacokinetic analysis

For the pharmacokinetic analysis of the myocardium and left ventricular blood, the time after contrast administration and contrast concentration were obtained for each voxel, and parameters were obtained by curve fitting using a two-compartment model (Figure 2c), applying Equations 2 and 3 for the myocardium and left ventricular blood pools, respectively. In this study, the CM was administered in two divided doses, and the concentration in blood or tissue at a given time is the sum of the pharmacokinetics of each dose. The model described by Brix et al and modified by Hoffman ^16–19^ was used to determine the concentration of CM in the myocardium (Ct).

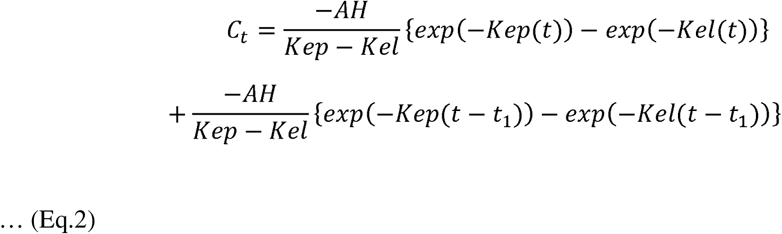

Where *K_el_* is the elimination constant of CM from the central compartment, *K_ep_* is the exchange rate constant from the extravascular extracellular space to the plasma, *AH* is the amplitude scaling constant in the equation modified by Hoffman and approximately corresponds to the size of the extravascular extracellular space ^19,20^

The CM concentration in the left ventricle (LV) blood pool (Cb) was calculated using the following bi-exponential equation ^9^:

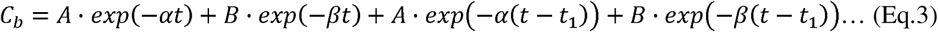

Where A and B are hybrid intercept terms, α and β are distribution and overall elimination rate constants. Where *t* is the time from the first injection, and *t_1_* is the time interval between 1^st^ and 2^nd^ injection. The contrast medium concentration at time t is calculated based on the principle of superposition with two doses^21^. Pharmacokinetic analysis was performed using the Python programming language (Python 3.9, Beaverton, Ore; https://www.python.org/). Parameters Ct and Cp were determined for each voxel. Fitting was performed using the curve_fit function, and optimization was performed using the Levenberg–Marquardt algorithm (Curve_fit function, Scipy 1.10.0). The source code for fitting is available online (https://github.com/yasuohta/dynamicT1map).

After voxel-wise pharmacokinetic (PK) analysis, measured concentration (MC) maps from T1 values and estimated concentration (EC) maps were simultaneously generated from the model for the myocardium and LV blood pool at the exact acquisition time of the post-CM T1 maps after CM administration. For all three cross-sections imaged, the difference between MC and EC of the myocardium or left ventricular pool was determined for each voxel, and the mean (delta_mean) and standard deviation (delta_SD) were calculated.

To determine the goodness-of-fit of the model, the coefficient of determination (R^2^) maps were calculated using following equation: 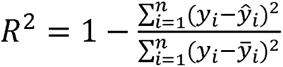, where y_i_ are the observed values, *ŷ_i_* the model−predicted values, and *y̅_*i*_* the mean of the observed values. This metric quantifies the proportion of total variance explained by the fit. The R^2^ value of the myocardial region was calculated for both the ER and non-registration datasets. Myocardium or LV blood pool regions were extracted using image masks (Supplementary Fig. S3).

### Statistical Analysis

Statistical analyses were performed using R statistical software (version 4.5.0; The R Foundation for Statistical Computing). The Shapiro–Wilk test was used to assess normality. Continuous variables were presented as mean ± SD or median and interquartile range for normally and non-normally distributed data, respectively. Categorical variables were expressed as absolute values and percentages. Linear regression analysis was performed to estimate the regression line and determine the predictive relationship between T1-derived concentration and reference. Comparisons of subjects’ backgrounds or goodness-of-fit (R^2^) were conducted using the t-test or Mann–Whitney U test (depending on normality) for continuous variables and the χ2 test or Fisher’s exact test for categorical variables. Model fit was compared across registration (no ER vs. with ER) and time points by R^2^ of measured on estimated median concentrations and reporting Akaike’s information criterion (AIC) and Bayesian information criterion (BIC). Prediction error (|measured – estimated|) was analyzed in R using the lme function with registration, time (factor), and their interaction as fixed effects and a random intercept per subject; analysis of variance (ANOVA) performed Type III tests of registration, time, and their interaction, and post hoc comparisons were conducted using paired t-tests with Bonferroni correction. The level of statistical significance was set at p < 0.05.

## Results

### Phantom study

Linear regression analysis yielded the following equation: T1-derived CM concentration = 1.01* (reference concentration) – 0.0273 (R^2^=0.999, p<0.001), confirming the linearity between the reference and the measured value within the range of contrast medium concentration up to 2.5mM/l in a phantom study (Fig. 3a). Bland-Altman analysis demonstrated the bias of 0.02mM/l (upper limit, 0.08mM/l, lower limit −0.04mM/l), (Fig. 3b).

**Fig. 3.**
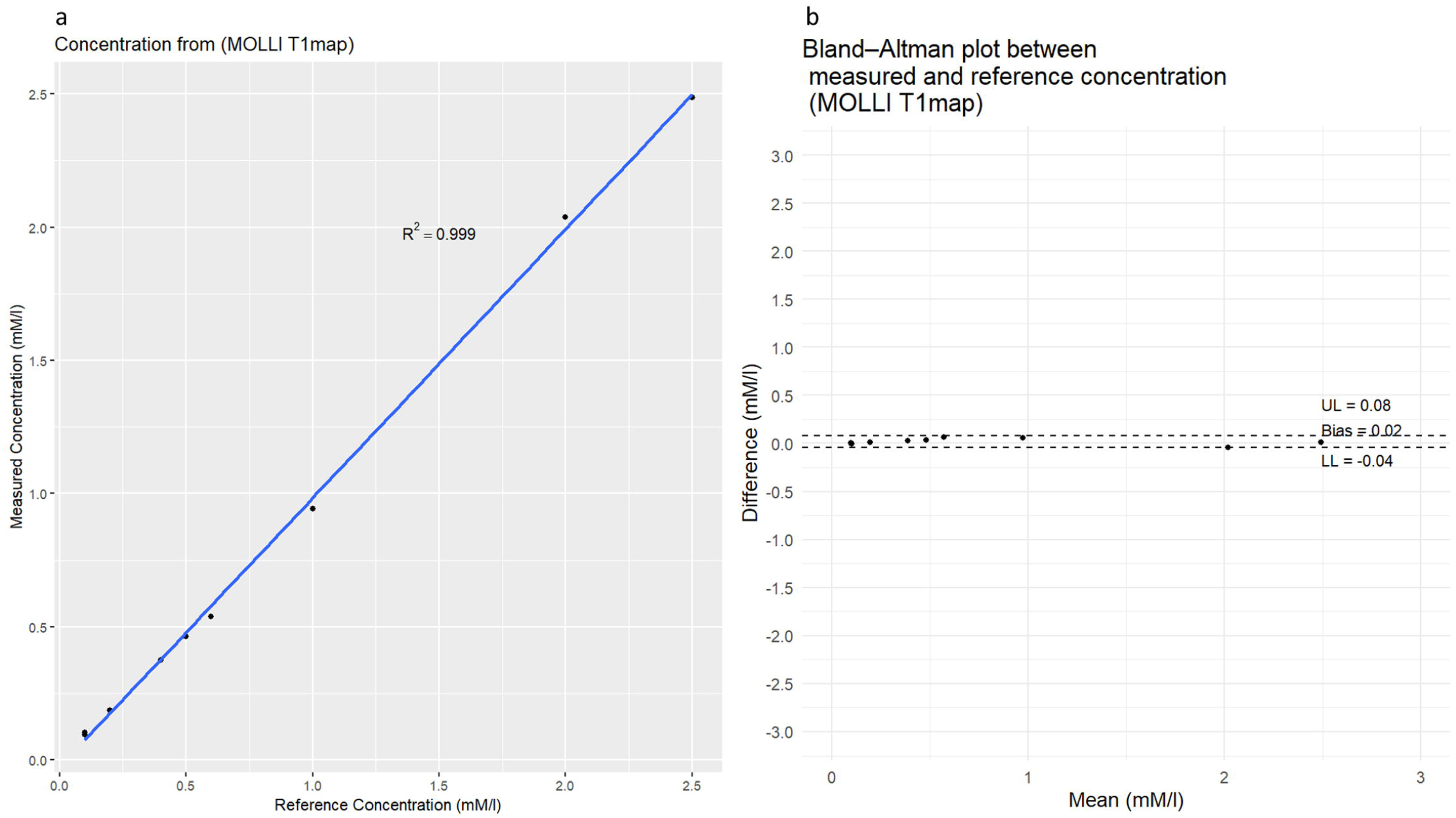
The regression analysis (a) and Bland-Altman plot (b) between diluted contrast medium concentration and measured gadobutrol concentration were calculated using T1 values measured by the MOLLI sequence. Correlation coefficient is R^2^=0.999 (p<0.001). Bland-Altman plot demonstrates a bias of 0.02mM/l (upper limit: 0.08mM/l, lower limit: −0.04 mM/l). *MOLLI* modified Look-locker, *UL* upper limit, *LL* lower limit

### In vivo evaluation

Analyzable dynamic T1 maps were obtained for all participants. The background characteristics of the participants at the time of the test request and the results measured by CMR are shown in Table 1.

**Table 1.**
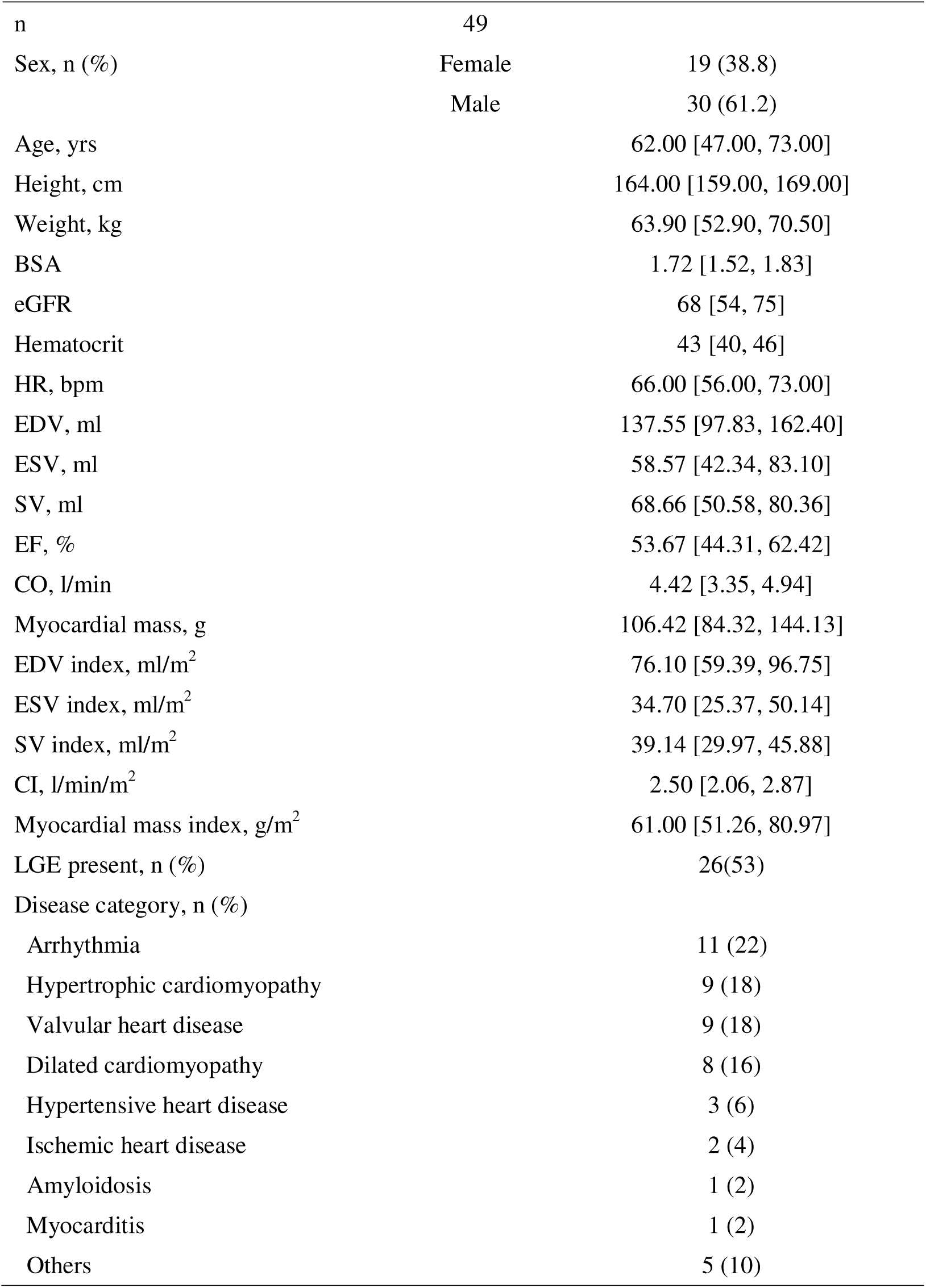
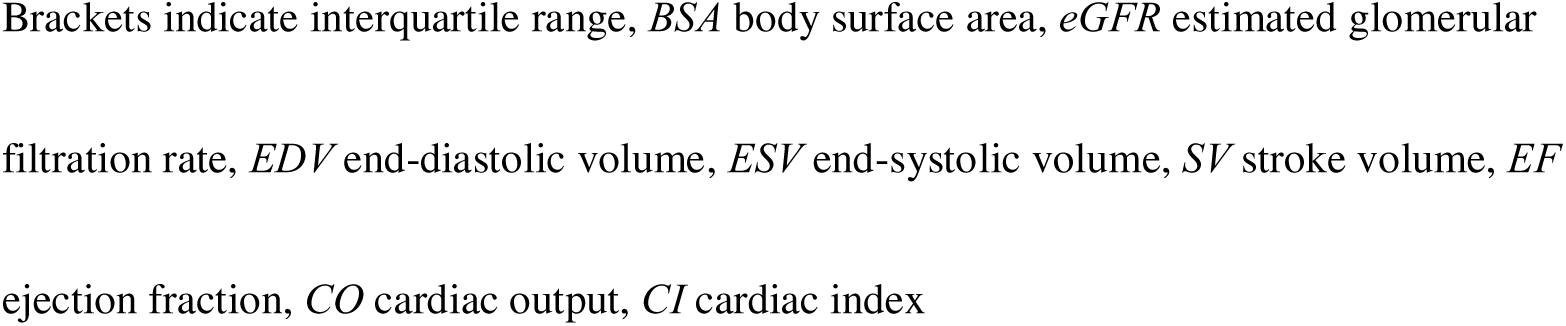
Participants’ backgrounds.

The concentration of CM in the myocardium decreased exponentially over time. A representative example is shown in Fig. 4. The mean CM concentrations in the myocardium with T1 map registration and without T1 map registration are presented in Fig. 5, and Table 2. Model fit, assessed by AIC and BIC for the regression of measured versus estimated median concentration at each time point, consistently favored the ER model over no ER at all four time points (ΔAIC ≤ –30, ΔBIC ≤ –30), indicating improved overall fit with ER. A repeatedlmeasures ANOVA on prediction error revealed a highly significant main effect of registration (p < .0001), indicating that co−registration reduced absolute error overall. Post-hoc pairwise comparisons showed that ER significantly lowered error at 5 min (p < 0.0001) and 9 min (p = 0.0155), but not at 2 min (p = 0.085) or 15 min (p = 0.343). Delta_mean was analyzed by repeated□measures ANOVA, which showed no overall effect of ER (p = 0.55), indicating that the influence of ER on delta_mean varied across measurement times. Detailed delta_mean values at each time point are shown in Table 2 and Fig. 5. Bland-Altman plot comparing measured and estimated CM concentration of myocardium is demonstrated in supplementary Fig. S1-5. Delta_SD was analyzed by repeated□measures ANOVA, which revealed a highly significant main effect of ER (p < 0.0001). Post-hoc paired t-tests demonstrated that ER significantly reduced delta_SD at every time point (each, p<0.001). Detailed delta_SDs are shown in Table 2.

**Fig. 4.**
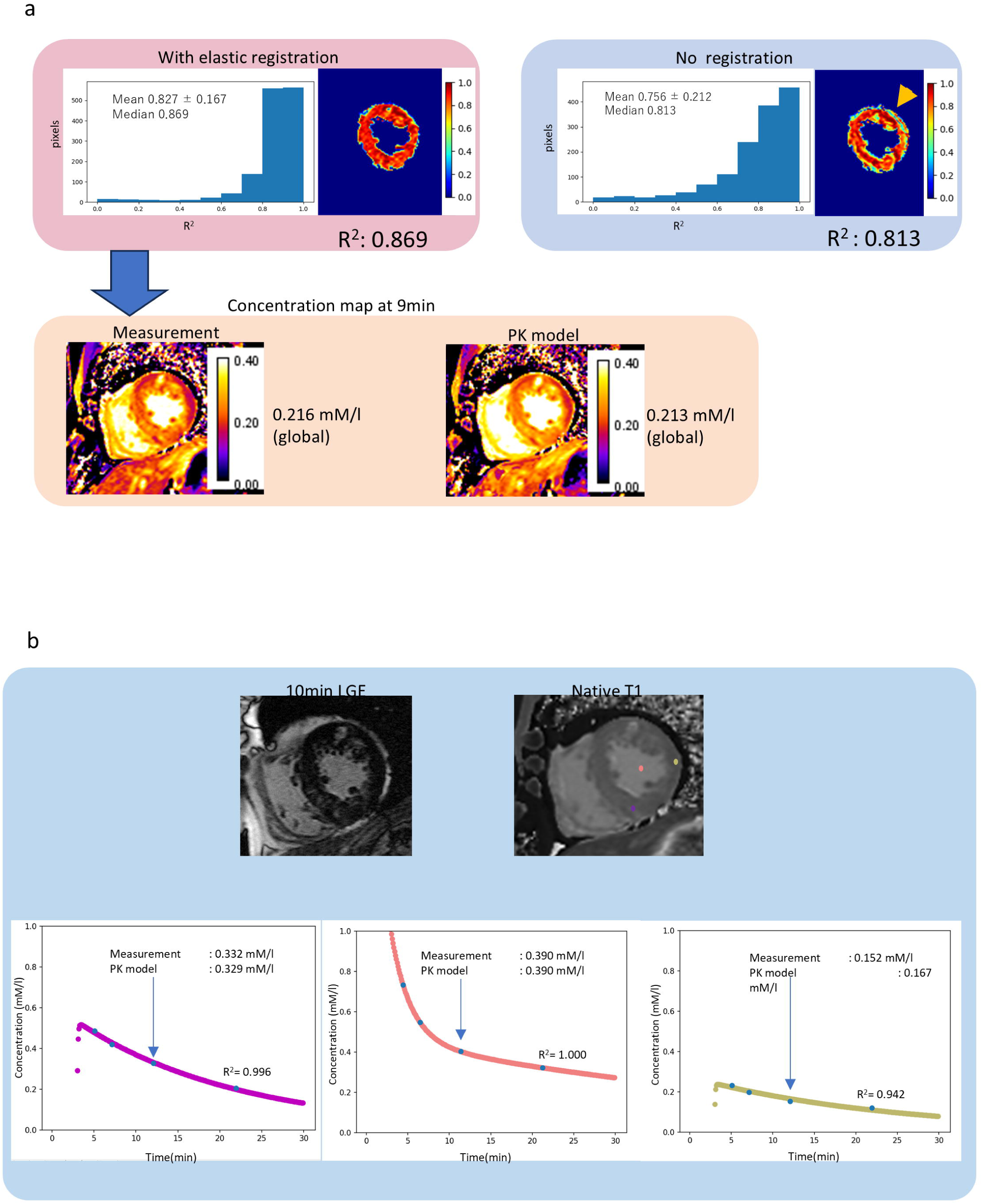
A representative example, a female with hypertrophic cardiomyopathy. **a** The coefficient of determination (R^2^) of myocardium from the PK. The mean R^2^ using the alignment on the upper right was 0.756±0.212, with a median of 0.813. The R^2^ map showed a deterioration in fitting at the myocardial edge (green-color-coded area). With the use of ER, the results on the upper left showed an improvement in the mean fitting coefficient of 0.827±0.167, with a median of 0.869, and a decrease in variability in the histogram. The R^2^ map with ER shows an increased red-colored area, which means a higher R^2^. The predicted myocardial average CM concentration (0.213 mM/l) at 9 min matches the concentration (0.216 mM/l) calculated from the actual T1 map. *PK* pharmacokinetics analysis, *ER* elastic registration. **b** The dots on the T1 map correspond to the region matching the LGE of the infero-septal wall (purple), the region without LGE in the lateral wall (yellow), and the left ventricular cavity (orange). The lower panel shows the time and concentration of CM injection in each region (blue dots) and the fitting curves by the PK model. The colored lines correspond to the region of the myocardium or LV blood pool indicated on the native T1 map. CM concentration values from the T1 measurement and PK model are indicated in each graph. *PK* pharmacokinetics analysis, *LGE* Late myocardial enhancement, *CM* contrast medium.

**Fig. 5.**
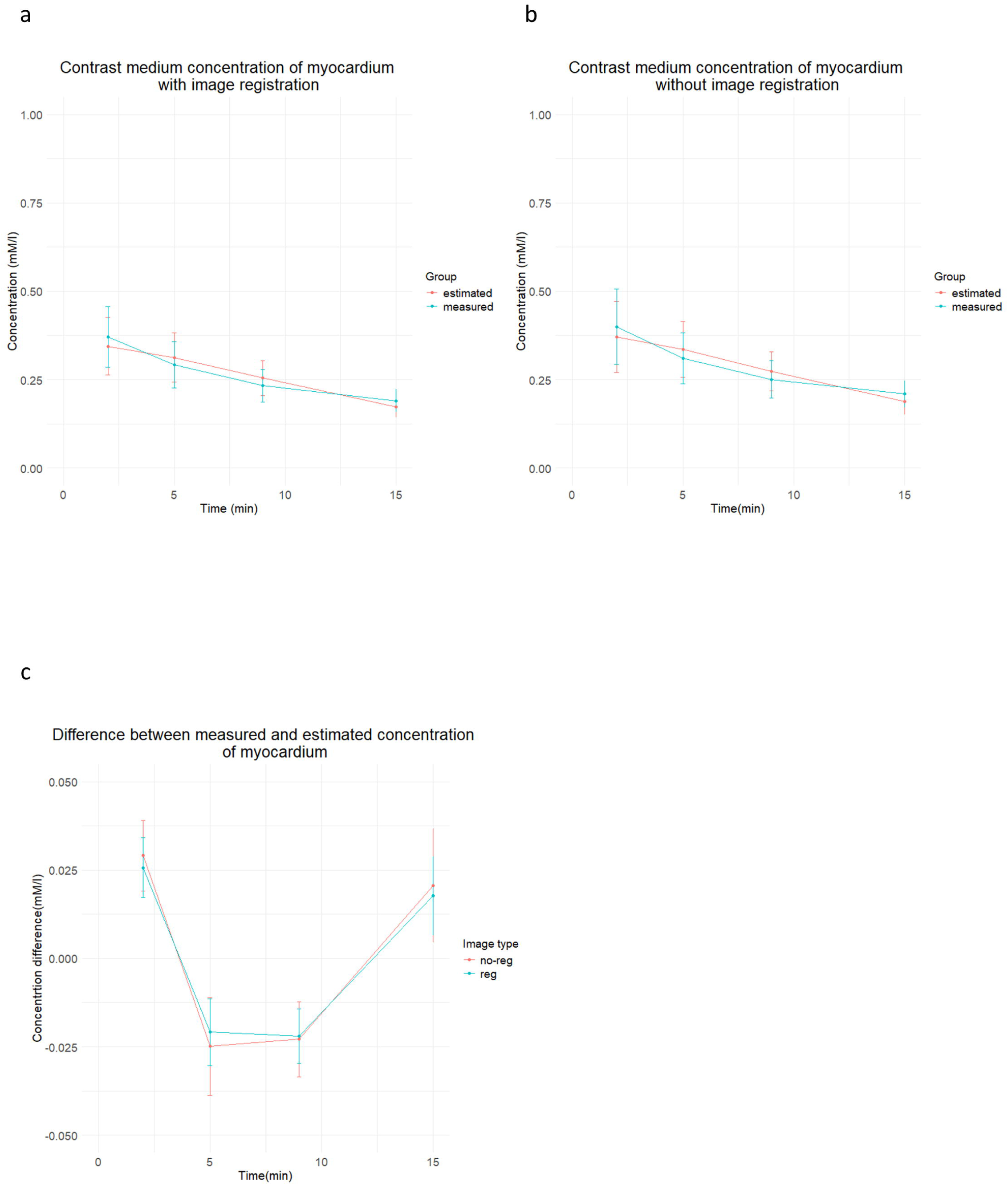
Contrast medium concentration of myocardium from measured T1 maps or estimated from PK model over time. Contrast medium concentration of myocardium from measured T1 maps or estimated from PK model over time, with ER (a) and without ER (b). The differences between measured and estimated contrast medium concentration of myocardium over time with (green line) and without (magenta line) ER (c). *PK* pharmacokinetic analysis, *ER* elastic registration, *no-reg* no registration, *reg* registration

**Table 2.**
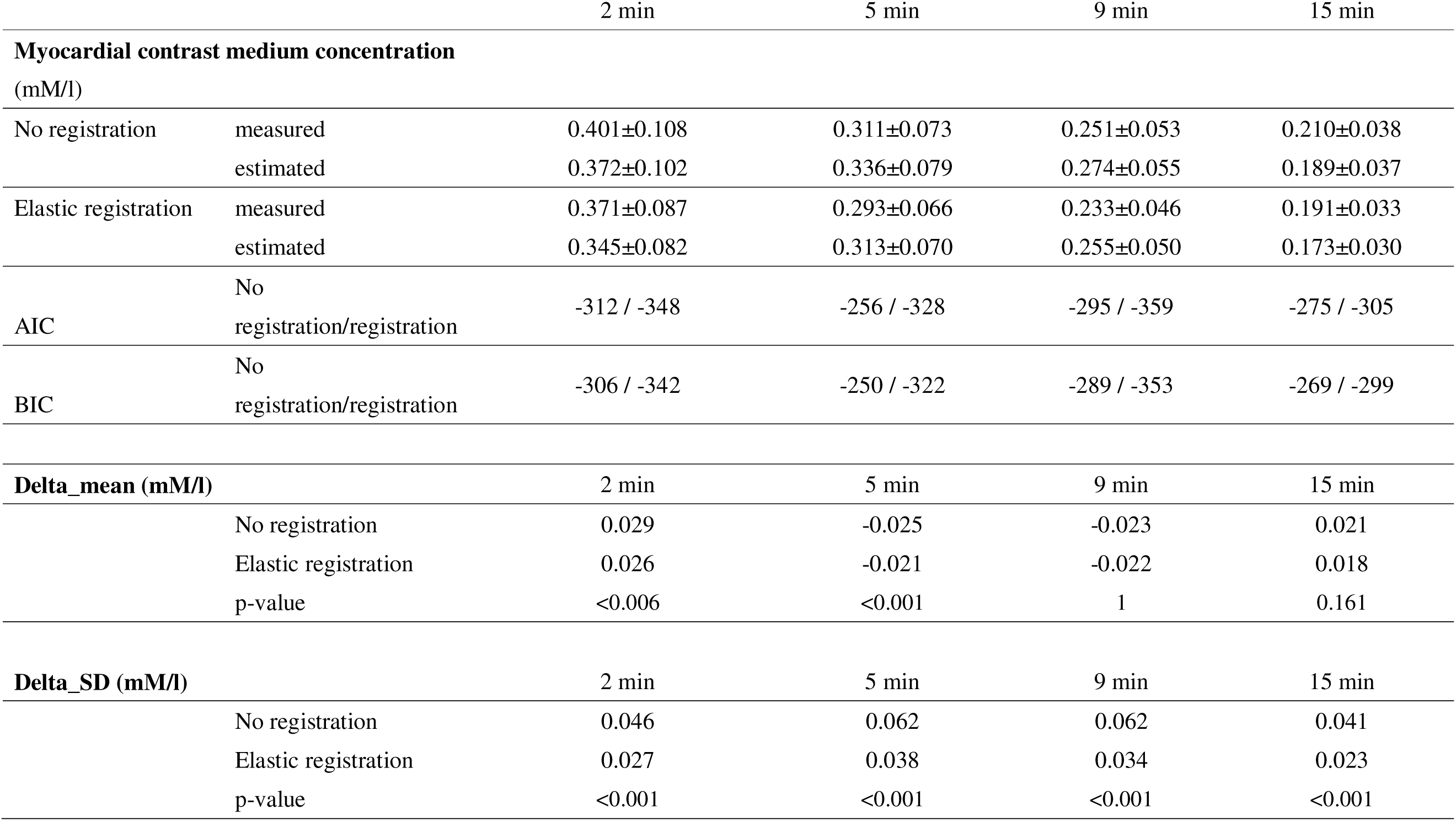
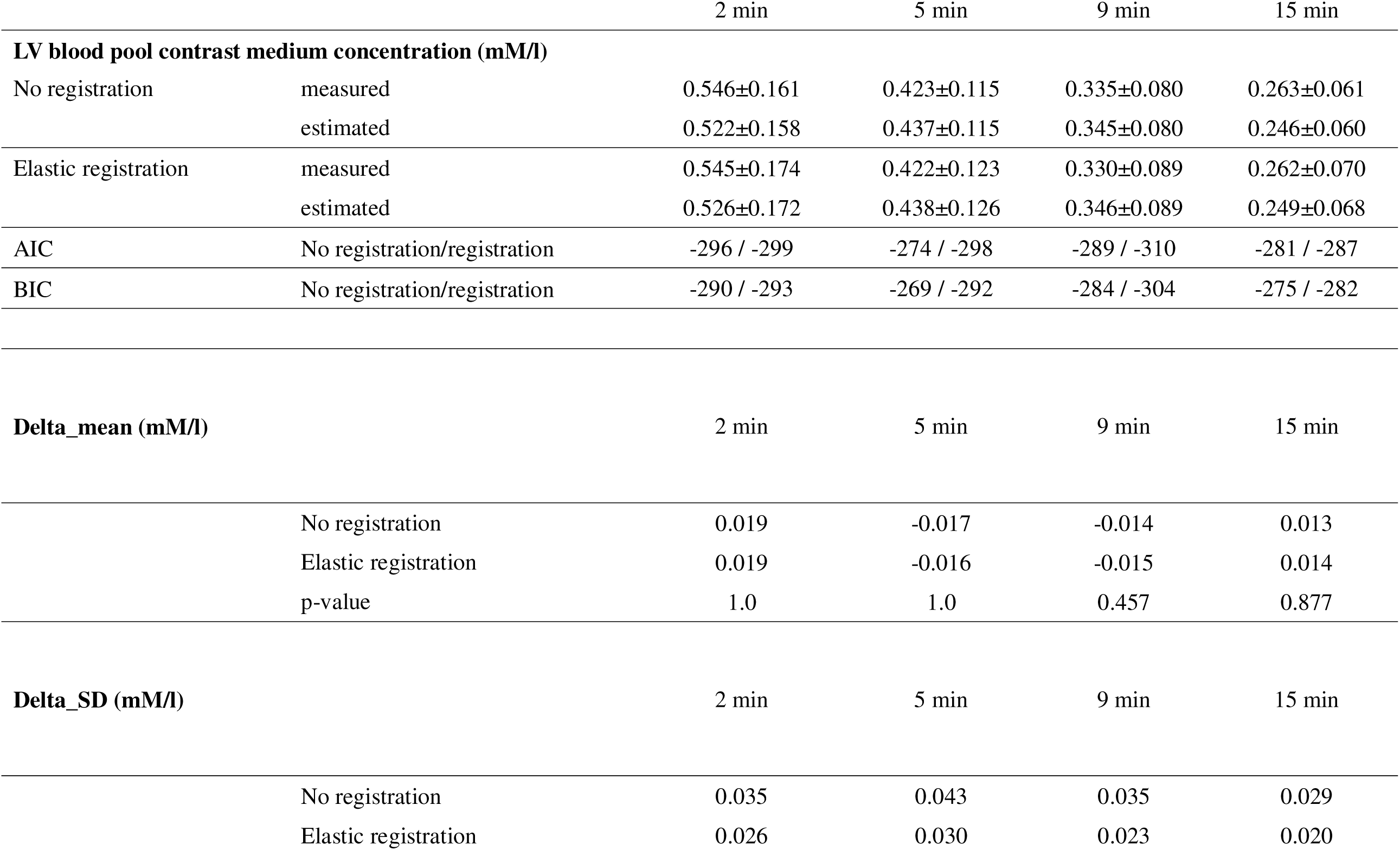
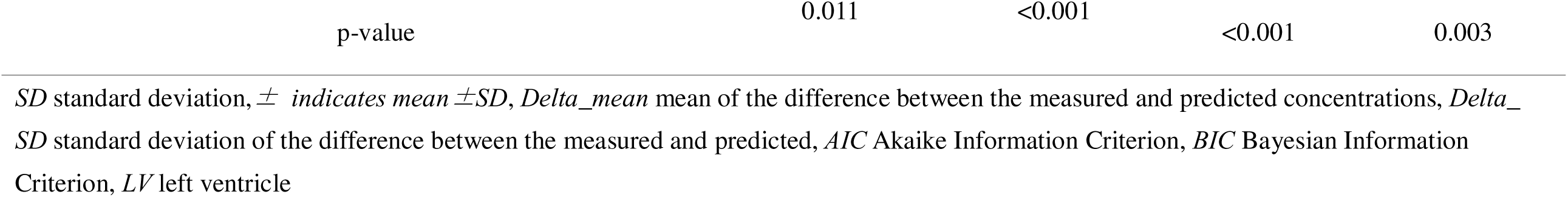
Myocardial and LV blood pool contrast medium concentration, difference, and SD between measured and estimated concentration with or without non-registration.

The CM concentration in the LV blood pool decreased exponentially over time. The mean CM concentrations in the myocardium with and without T1 map registration are presented in Fig. 6, and Table 2. Model fit, assessed by AIC and BIC for the regression of measured versus estimated median concentration at each time point, consistently favored the ER model over no ER at all four time points (ΔAIC ≤ –3, ΔBIC ≤ –3), indicating improved overall fit with ER. A repeated□measures ANOVA revealed no main effect of ER (p = 0.37). Post-hoc pairwise comparisons confirmed that no significant differences were found at any time point (all, p > 0.30). Delta_mean was analyzed by repeated□measures ANOVA, which revealed no main effect of ER (p = 0.71). Registration did not significantly alter delta_mean at any individual time point (Table 2). Detailed delta_mean values at each time point are shown in Table 2 and Fig. 6. Bland-Altman plot comparing measured and estimated CM concentration of LV is demonstrated in supplementary Fig. S6-9. The delta_SD was analyzed by repeated□measures ANOVA, which revealed a highly significant main effect of ER (p < 0.0001). Post□hoc paired t-tests showed that ER significantly reduced delta_SD at every time point. Detailed delta_SDs are shown in Table 2.

**Fig. 6.**
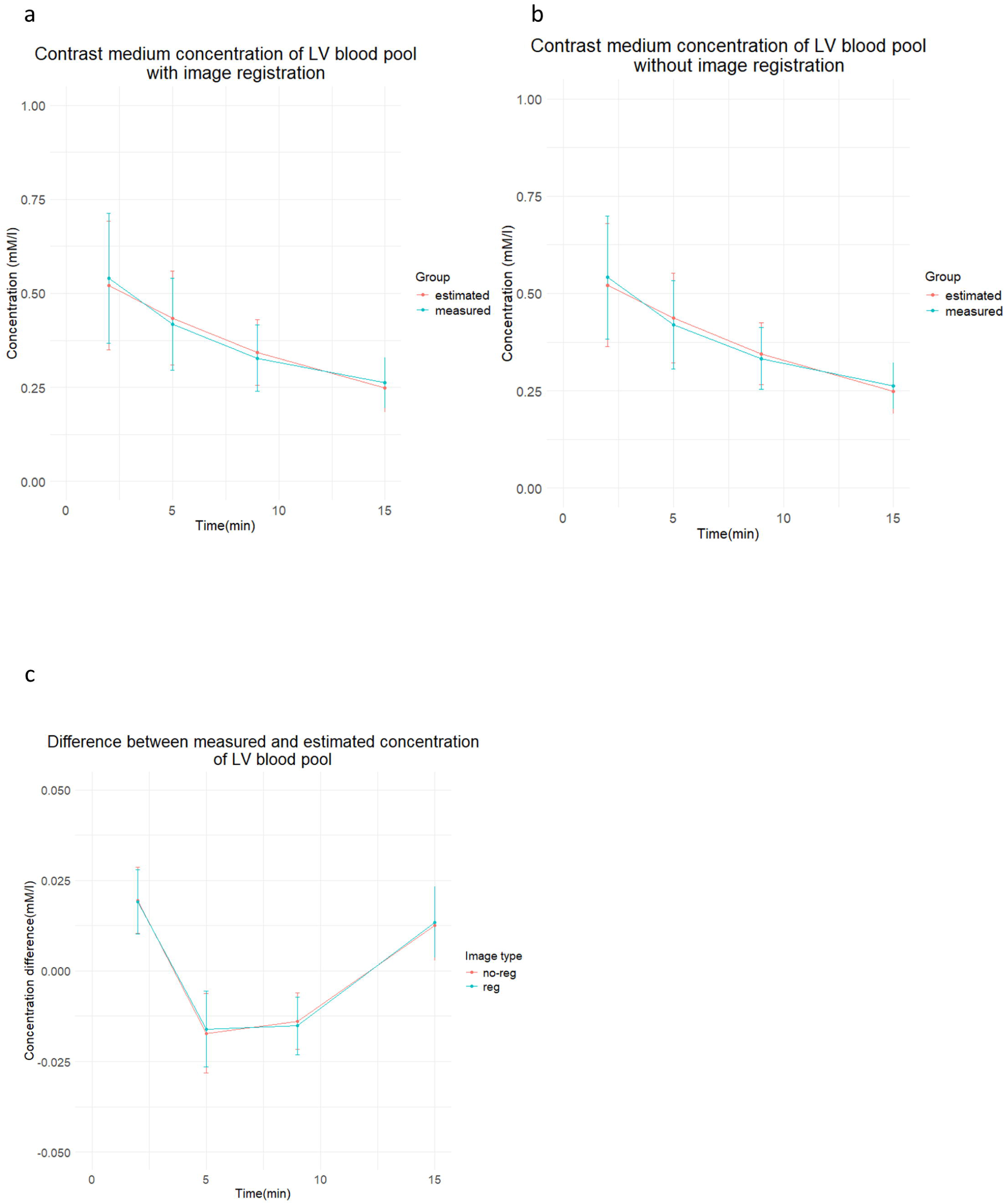
Contrast medium concentration of LV blood pool from measured T1 maps or estimated from PK model over time. Contrast medium concentration of left ventricular blood pool from measured T1 maps or estimated from PK model over time, with ER (a), without ER (b). The differences between measured and estimated contrast medium concentration of left ventricular blood pool over time, with ER (green line), without ER (magenta) (c) *PK* pharmacokinetic analysis, *ER* elastic registration, *no-reg* no registration, *reg* registration

The R^2^ in the myocardial CM concentration analysis improved significantly from 0.795 to 0.816 when ER was applied (p = 0.004). The R^2^ for the blood pool improved significantly from 0.847 to 0.902 when the ER was applied (p < 0.001) (Table 3).

**Table 3.**
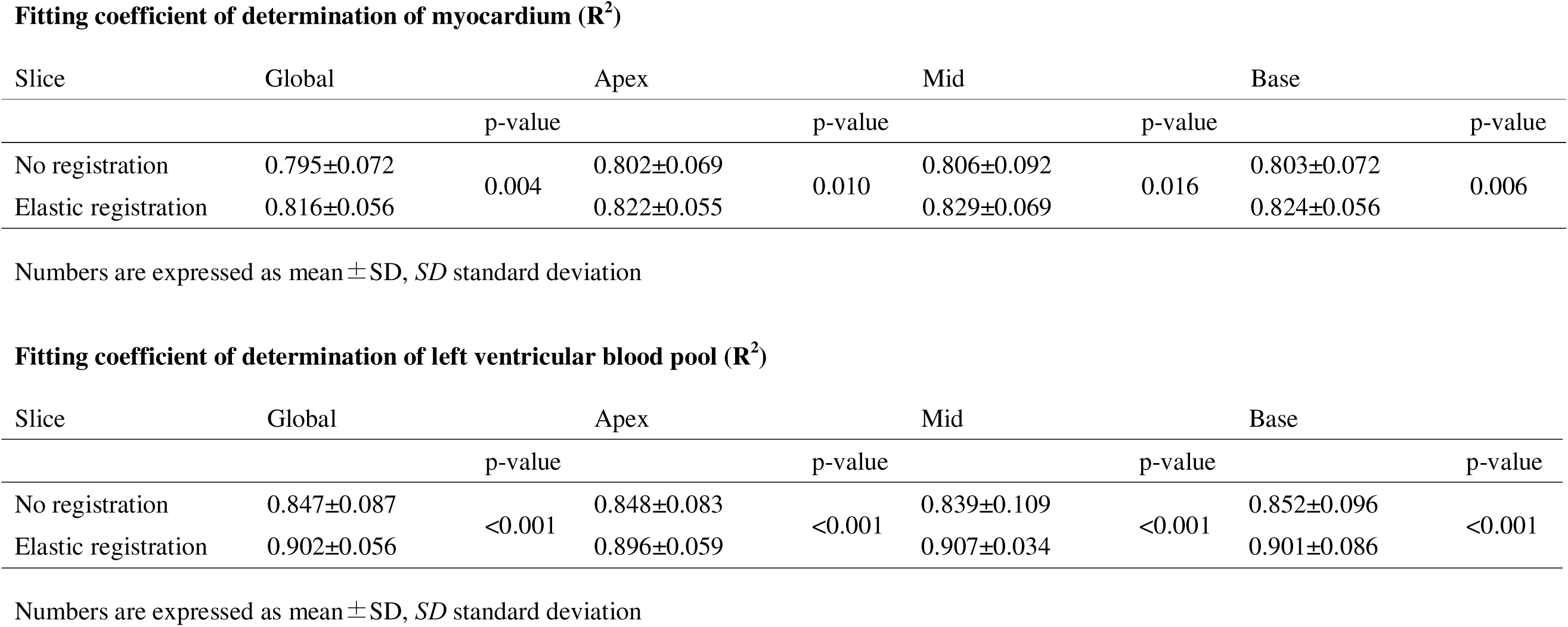
Fitting coefficient of determination of myocardium (R. ^2^)

| Slice | Global |  | Apex |  | Mid |  | Base |  |
| --- | --- | --- | --- | --- | --- | --- | --- | --- |
|  |  | p-value |  | p-value |  | p-value |  | p-value |
| No registration | 0.795±0.072 | 0.004 | 0.802±0.069 | 0.010 | 0.806±0.092 | 0.016 | 0.803±0.072 | 0.006 |
| Elastic registration | 0.816±0.056 |  | 0.822±0.055 |  | 0.829±0.069 |  | 0.824±0.056 |  |

**Fitting coefficient of determination of left ventricular blood pool ( $R^2$ )**
| Slice | Global |  | Apex |  | Mid |  | Base |  |
| --- | --- | --- | --- | --- | --- | --- | --- | --- |
|  |  | p-value |  | p-value |  | p-value |  | p-value |
| No registration | 0.847±0.087 | <0.001 | 0.848±0.083 | <0.001 | 0.839±0.109 | <0.001 | 0.852±0.096 | <0.001 |
| Elastic registration | 0.902±0.056 |  | 0.896±0.059 |  | 0.907±0.034 |  | 0.901±0.086 |  |

Numbers are expressed as mean ± SD, *SD* standard deviation

## 4. Discussion

This study demonstrates that the MOLLI method enables the generation of CM concentration maps over time, predictive concentration maps can be generated from pharmacokinetic analysis models, and that the goodness-of-fit improves when ER are used.

The use of ER ^22^ to fit each image post-contrast to the native T1 map was a key technique for pharmacokinetic analysis in this study, enabling concentration mapping and voxel-by-voxel pharmacokinetic analysis, improving the goodness-of-fit, and reducing the difference between the measured and predicted values. So far, the alignment of images acquired with MOLLI for T1 map creation has enabled the creation of T1 maps without misalignment ^3,4,23^ and has been widely applied in clinical practice. Furthermore, in ECV map creation, the alignment between pre- and post-contrast maps has been improved during imaging or after imaging [21–23]. In this study, image co-registration was easily carried out using the workstation’s functions.

Tissue CM concentration calculations have mainly been used to measure the permeability of blood vessels in tumors, assess tissue properties, and determine efficacy after treatment ^24–26^. In the myocardium, first-pass perfusion assessment is often used to diagnose ischemia; however, tissue assessment by observing washout after contrast is limited ^27–29^. A previous study evaluated CM kinetics in amyloidosis using T1 values ^30^; however, unlike CM concentrations, post-contrast T1 values are influenced by both CM concentrations and native T1 values, which poses a challenge for quantitative evaluation using post-contrast T1 values. Nonetheless, kinetic parameters can be analyzed using this approach as new parameters for lesion characterization and as predictors of CM concentration at arbitrary times. Although this approach requires the acquisition of native T1 values and image alignment among images after contrast, dynamic contrast-enhanced (DCE)-MRI using two-flip angles with an ultrashort echo time has been reported to provide accurate contrast density measurements without pre-contrast scanning ^31^ and has been used in the same way in the case of MRI with a two-flip angle, with high expectations for the future. On the contrary, this T1 map-based approach applies to many types of clinically available equipment. In pharmacokinetic analysis, various modeling approaches have been proposed. In the present study, we adopted the Brix model, which has been commonly used in tissue evaluations. While several alternative models ^20,32^ have been reported, the limited number of temporal sampling points in our dataset led us to choose the Brix model, considering the number of parameters required for fitting. For instance, the Tofts model typically requires a relatively long dynamic acquisition with approximately 20 time points and arterial input function ^20^, and was therefore not applied in this study. However, recent advancements in myocardial perfusion imaging, such as the use of dual-sequence techniques for quantitative MBF assessment ^33,34^, have enabled more precise evaluation of contrast dynamics using perfusion sequences. Application of such models to myocardial tissue is expected to be increasingly feasible in the future.

In the present study, the CM was administered in two separate injections. This was done for two main reasons. First, the protocol was designed to accommodate split dosing as in stress perfusion imaging. Second, divided administration reduced the initial peak concentration of CM in the cardiac chambers compared to single-dose injection, thus maintaining linearity between the T1 values and CM concentrations within a safe dynamic range. Even with split dosing, contrast concentration can be estimated as the sum of concentrations following each administration time point, based on the principle of superposition^21^. While the amplitude of the concentration-time curve may differ from that of a single bolus injection, the inflow and outflow rate constants theoretically remain unchanged. Therefore, pharmacokinetic parameters can be estimated independently of injection interval, supporting the robustness and potential standardization of the derived indices.

Late myocardial enhancement is used for assessment of myocardial properties, has shown a high correlation with myocardial fibrosis ^1,2,35^, and is a major motivation for performing cardiac MRI for tissue assessment ^36,37^. However, the evaluation is only qualitative or semi-quantitative, with remote normal myocardium as reference, and the time after contrast administration must also be precisely matched for comparison. Quantitative values derived from cardiac MRI, such as T1 mapping and ECV, are now used to detect diffuse fibrosis and subtle changes that are difficult to detect using LGE ^38^. Due to the differences in values between devices and sequences, it is recommended to have standard values at each facility [39, 40], with a standardized approach that transcends facilities and equipment. Although ECVs show limited variation across devices, standardizing the exact imaging time after CM administration for large-scale comparisons is impractical, and measuring hematocrit is also necessary. In this context, the proposed method could serve as a standardized index because it provides a consistent analytical scale based on concentration values and enables the creation of concentration maps at any post-imaging time. This promotes uniform assessment and supports longitudinal studies. Clinically, this approach enables the simultaneous quantification of CM concentration in both the myocardium and blood pool, supporting ongoing evaluation of contrast kinetics, including disease-specific enhancement and washout patterns. By combining synthetic LGE images from post-contrast T1 values [42, 43] and time-aligned T1 maps from the PK model, accurate synthetic LGE images synchronized with the post-CM administration time can be generated. Variability caused by disease type, sex, imaging sequence, and contrast dose needs further investigation in large cohorts to establish reference values for clinical use. The proposed method showed improved pixel-wise fitting, supporting its potential for detailed regional analysis, including segmental evaluation and comparisons based on LGE presence. Although LGE-positive cases were included, analyses of contrast differences between fibrotic and normal myocardium were not performed and warrant future investigation.

## 5. Limitations

This study has limitations. First, this study was conducted with a single scanner at a single site and should be evaluated using images from other manufacturers. Second, curve fitting may be affected by the time and frequency of dynamic imaging. Although late post-contrast T1 mapping may improve prediction accuracy, clinical constraints limited imaging to an earlier, feasible time point. For this reason, the model proposed by Brix et al., with a small number of parameters, was used. The optimal number of fitting points and models should be studied in the future. Third, we did not compare the contrast concentration estimated by curve fitting with the actual gadolinium CM concentration. Since there is no standard method to precisely measure the contrast medium (CM) concentration in the myocardium, this study’s CM concentration reference is based on a T1 map calculation. Its reliability should be validated against other approaches, like the Tofts model or directly comparing estimated blood concentration and actual blood CM concentrations. Although direct blood measurements were not feasible here, phantom studies have demonstrated a strong correlation between T1-derived and known CM concentrations when using the MOLLI method. The National Library of Medicine site also reports a concentration in plasma of 0.441 mM/l [range: 0.281,0.829] after 20 min at a single dose of 0.1 mmol/kg in healthy adults ^39^. The predicted concentration of 0.316 mM/l in the blood pool at 20 min in our cohort was slightly above normal, with a plasma concentration of 0.526 mM/l at 40% hematocrit. However, the study included patients with moderate or severe renal dysfunction, and the half-life of gadobutrol was prolonged in patients with reduced renal function ^40^, which may be influenced by differences in the background of the participants. In this study, CM concentrations in whole blood and myocardial CM concentrations are evaluated. Although CM concentrations in blood and plasma can be evaluated between individuals after conversion by hematocrit, the extent to which hematocrit values in small intramyocardial vessels differ between individuals and their impact on myocardial CM concentrations need to be evaluated.

## 6. Conclusion

In conclusion, ER of the T1 map taken over time after contrast, combined with the native T1 map, improves voxel-wise fitting through pharmacokinetic analysis of the myocardium and blood pool. It also reduces the gap between measured values and predictions from pharmacokinetic analysis models. Concentration provides a simple metric for comparison. Because of its usefulness in longitudinal assessments, this approach has the potential to become a standard measure for myocardial evaluation, provided it is validated across different scanners and imaging protocols.

## Declaration of generative AI and AI-assisted technologies in the writing process

We declare that we have not used any generative AI or AI-assisted technologies in the writing process of this work.

## Acknowledgment

We would like to thank Editage (www.editage.jp) for English language editing.

## Data availability

The datasets for this study are not publically available due to Japanese data protection laws. The data that support the findings of this study are available from the corresponding author upon reasonable request.

## Author contributions

Yasutoshi Ohta: Writing – original draft, Visualization, Validation, Methodology, Investigation, Software, Funding acquisition, Formal analysis, Data curation

Masaru Shiotani: Writing – review & editing, Validation, Data curation,

Yoshiaki Morita: Writing – review & editing, Validation

Tatsuya Nishii: Writing – review & editing, Validation

Hiroki Horinouchi: Writing – review & editing,

Akiyuki Kotoku: Writing – review & editing,

Midori Fukuyama: Writing – review & editing,

Emi Tateishi: Writing – review & editing,

Tetsuya Fukuda: Writing – review & editing, Supervision,

## Additional information

Authors declare no conflicts of interest.

This work was supported by JSPS KAKENHI (No. 22K15835).

