## Supplemental material for "Estimation of Myocardial and Blood Gadolinium Concentrations from T1 Mapping via Pharmacokinetic Modeling: Influence of Elastic Deformation Registration"

**Supplementary material**

**Phantom study**

The phantom was used to confirm the linearity and correlation between the diluted CM concentration and the CM concentrations calculated from T1 values.　The CM (gadobutrol; Gadovist®, Bayer-Yakuhin, Osaka, Japan) was diluted in normal saline to the following concentrations (0.1, 0.2, 0.4, 0.5, 0.6, 1.0, 2.0, and 2.5 mM/l) and stored in 15ml vials. A phantom (Supplementary Fig. 1) was created by surrounding the vials with normal saline. The phantom was imaged using a 3T MRI system (MAGNETOM Vida; Siemens Healthcare, Erlangen, Germany) and a 30-channel body coil. T1 mapping was performed using a motion-corrected Look-Locker inversion recovery (MOLLI) sequence with 5 (3)3 and 4(1)3(1)2 acquisition schemes, for nativeT1map and post-contrast T1map [1, 2], respectively. The imaging parameters used were: TR/TE/flip angle, 3.8 msec/1.2 msec/35; slice thickness, 8 mm; voxel size, 1.4 ×1.4 ×8.0 mm^3^, NEX 1, FOV 360 x 306, matrix 256 x 144, parallel processing factor 2, initial effective inversion time 100msec with an increment of 80msec. ECG gating was used with a simulated ECG at 60 bpm from the system function. After imaging the vials at the center level of the phantom, T1 maps were automatically generated by the system. Circular regions of interest (19mm^2^) were placed at the center of the vials on images, and the T1 values for each concentration were measured. The native T1 value of normal saline was measured using MOLLI for native T1 acquisition, and the post-contrast MOLLI scheme was used for the T1 value measurement of contrast medium-contained vials. The contrast medium concentration was then calculated using equation (Eq.1).

$\frac{1}{T_{1_{p}}}=r_{1}c+\frac{1}{T_{1_{0}}}$ …（Eq.1）

where $T_{1_{ｐ}}$ indicates the post-contrast T1 value of each time point, r_1_ indicates the relaxivity value of gadobutrol at 3.0 Tesla, c indicates concentration of contrast medium (mM/L), $T_{1_{0}}$ indicates native T1 value. The r_1_ value for gadobutrol varies slightly between reports[3], and the r_1_ value of 5.0 given in the product monograph or report by the American College of Radiology was used [4, 5].

An almost perfect correlation (r=0.999, P<.001) between the diluted CM concentration and the CM concentration measured using MOLLI was demonstrated using a phantom study, confirming the linearity between the index and the measured value within the range of contrast medium concentration up to 2.5mM/l. Measured concentration by MOLLI = 1.01* (reference concentration) – 0.0273 (Pearson’s r = 0.999, p<.001). Bland-Altman analysis demonstrated the bias of 0.02mM/l (upper limit, 0.08mM/l, lower limit -0.04mM/l) .

**Pharmacokinetic analysis**

For pharmacokinetic analysis of myocardium and left ventricular blood, time after contrast administration and contrast concentration were obtained for each pixel, and parameters were obtained by curve fitting using a two-compartment model, applying Equations 2 and 3 for the myocardium and blood pools, respectively. The contrast medium concentration in the myocardium was calculated using the following equation from Brix et al. [6, 7] (Eq. 2).

…(Eq.2)

$$C_{t}=\frac{-AH}{Kep-Kel}\left\{ exp\left( -Kep(t) \right)-exp\left( -Kel(t) \right) \right\}$$

C_t_:	concentration of contrast medium (CM) in tissue

K_el_:	the elimination constant of the CM from the central compartment

K_ep_:	exchange rate constant from the EES to plasma

AH:	amplitude scaling constant

Contrast medium concentration in LV blood pool was calculated by the following equation (Eq. 3)

$C_{b}=A*exp\left( -\alpha t \right)+B*exp\left( -\beta t \right)$	… (Eq. 3)

A and B are hybrid intercept terms, α and β are distribution and overall elimination rate constants.

C_b_:	concentration of contrast medium (CM) in blood pool

V_p_:	distribution volume in plasma

A:	D(K_ep_ -α)/ V_p_(β-α)

B:	D(K_ep_ -β)/V_p_(α-β)

D:	contrast medium dose

α:	(K_el_+K_12_+K_ep_+z)/2

β:	(K_el_+K_12_+K_ep_-z)/2

z:	{(K_el_+K_12_+K_ep_)^2^-4･k_ep_ ･ K_el_ }^1/2^

As the contrast medium was administered in two divided doses, the contrast medium concentration Ct (Eq. 2) and the contrast medium concentration Cp (Eq. 3) in the blood pool after the completion of the two doses can be calculated using the following equations, assuming that the administration interval is t_1_.…（Eq 2‘）

$$C_{t}=\frac{-AH}{Kep-Kel}\left\{ exp\left( -Kep(t) \right)-exp\left( -Kel(t) \right) \right\}+\frac{-AH}{Kep-Kel}\left\{ exp\left( -Kep(t-t_{1}) \right)-exp\left( -Kel(t-t_{1}) \right) \right\}$$

And

$C_{p}=Aexp\left( -\alpha t \right)+Bexp\left( -\beta t \right)+Aexp\left( -\alpha\left( t-t_{1} \right) \right)+Bexp\left( -\beta\left( t-t_{1} \right) \right)$　…(Eq 3’)

Pharmacokinetic analysis was performed using the Python programming language (Python 3.9, Beaverton, Ore; https://www.python. org/). Parameters for Ct and Cp were determined for each pixel. Fitting was performed using the cureve_fit function and optimization using the Levenberg-Marquardt algorithm (cureve_fit function, Scipy 1.10.0).

The time of contrast administration was defined as the time at which the contrast medium reached the left ventricle after checking each perfusion image. The time was obtained from the Digital Imaging and Communications in Medicine (DICOM) header. The acquisition time of the post-contrast T1 map was obtained from the DICOM tag for each image, and the acquisition time of the first image of the MOLLI scheme was used for actual imaging time.

To determine the model's fitting accuracy, the coefficient of determination (R^2^) was calculated after calculating the residuals using the equation (Eq. 4) below.

$R^{2}=1-\frac{\sum_{i=1}^{n} {(y_{i}-\hat{y}_{i})}^{2}}{\sum_{i=1}^{n} {(y_{i}-\bar{y}_{i})}^{2}}\boldsymbol{}$… (Eq. 4)

where *i* are each pixel, y_i_ are the observed values, $\hat{y}_{i}$the model‐predicted values, and $\bar{y}_{i}$ the mean of the observed values. The R^2^ was calculated for each pixel, and an R^2^map was created.

**Masked image preparation:**

For the masked images of myocardium, regions within the threshold (1000 msec to 1400 msec) were extracted as myocardium after removing non-cardiac regions from the native T1 map.

As native T1 value of blood ranges was demonstrated as 1600-2200 msec at 3T [8, 9], the left ventricular blood pool was similarly depicted using threshold values. The mask image was confirmed by visual assessment and the threshold was fine-tuned if the mask did not match the myocardium or blood pool. Image J (National Institute of Health, Bethesda, MA) was used to create the mask. Contrast medium concentration or R^2^ map of the myocardium or left ventricular blood pool was extracted by applying each mask image (Supplementary Fig. S1).

**Supplementary Fig. S1**

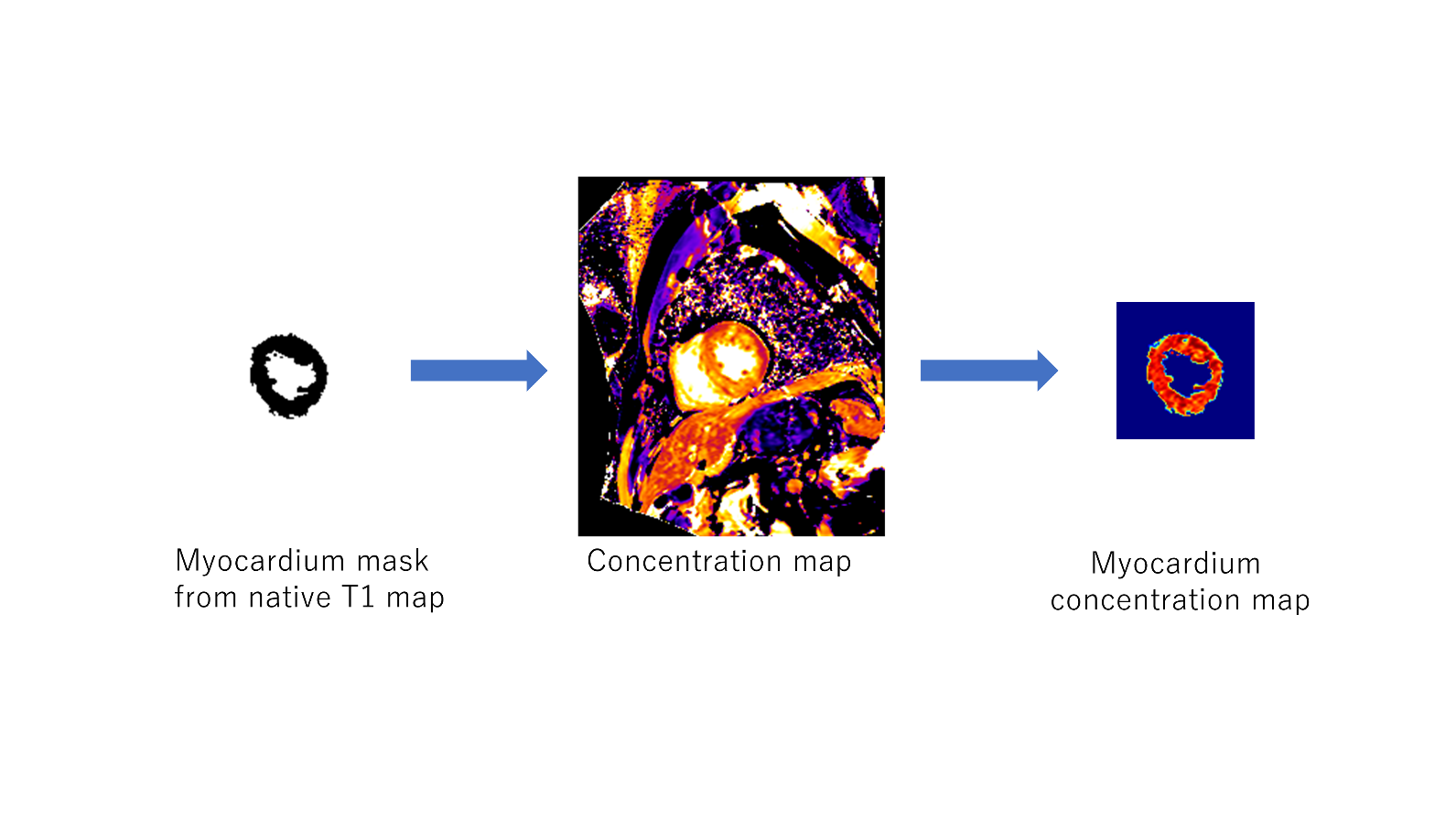

An example of myocardial density map extraction: a mask image of the myocardium or left ventricular blood pool is created from the Native T1 map, and a mask process is performed on the concentration map to extract a contrast medium concentration map of the myocardial (or left ventricular blood pool) region only.

**Supplementary Fig. S2**

Bland-Altman plot comparing measured and estimated contrast medium concentration of myocardium with (upper) or without (bottom) image co-registration, 2 minutes after injection. *CM* contrast medium

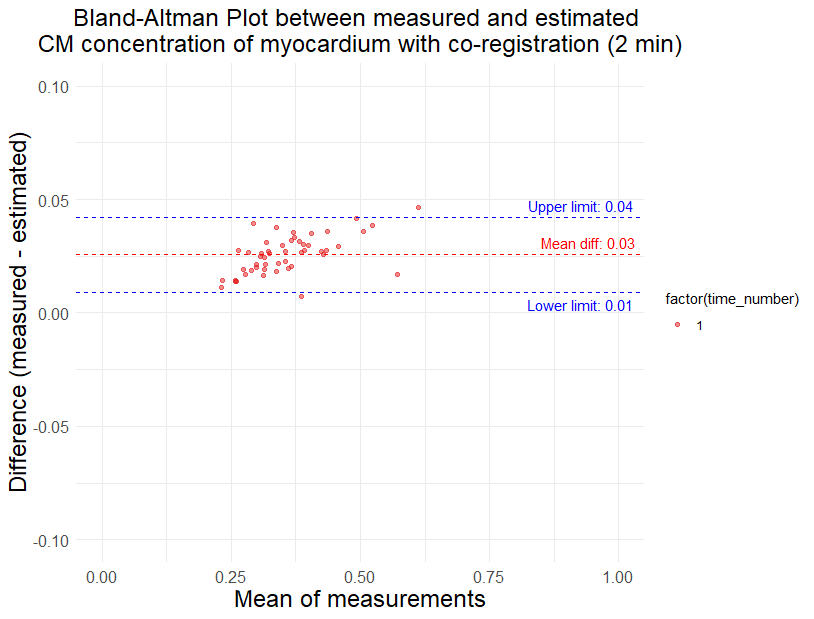

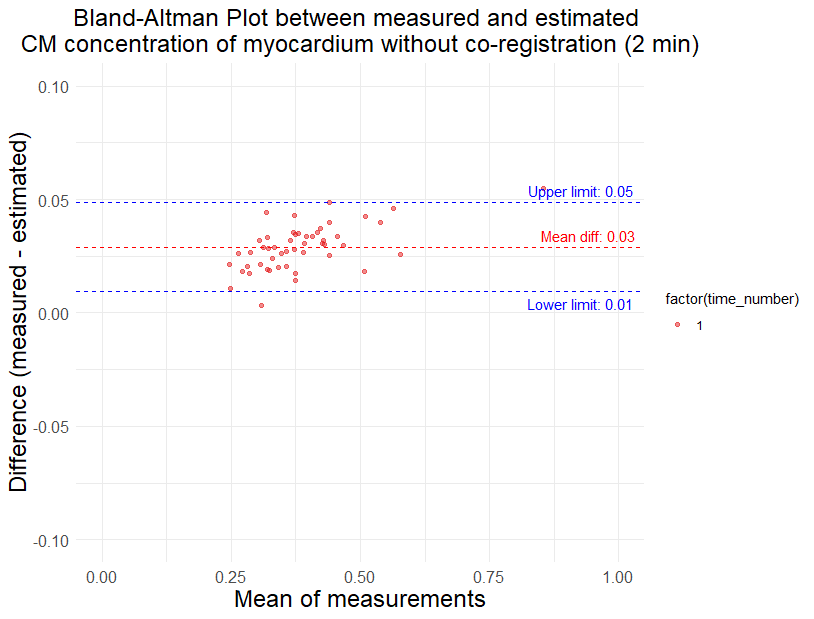

**Supplementary Fig. S3**

Bland-Altman plot comparing measured and estimated contrast medium concentration of myocardium with (upper) or without (bottom) image co-registration, 5 minutes after injection. *CM* contrast medium

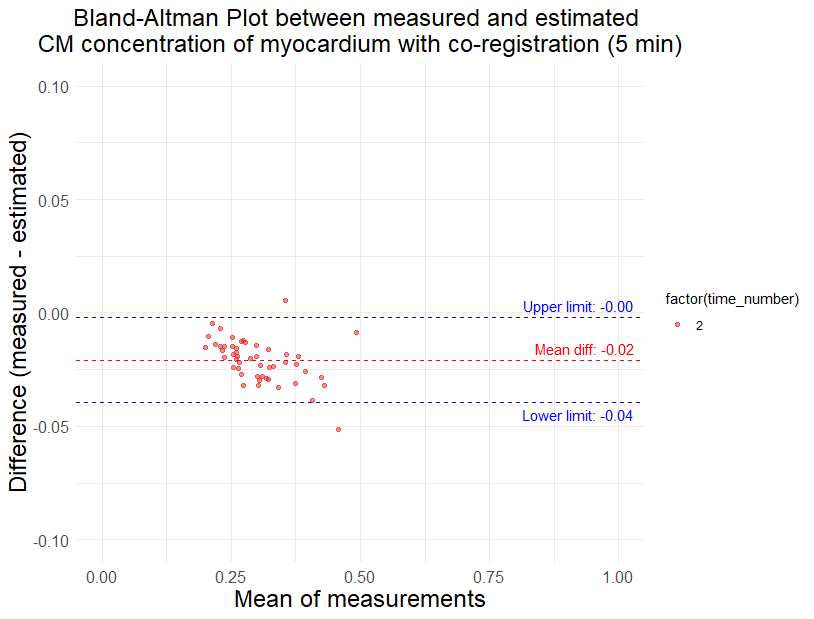

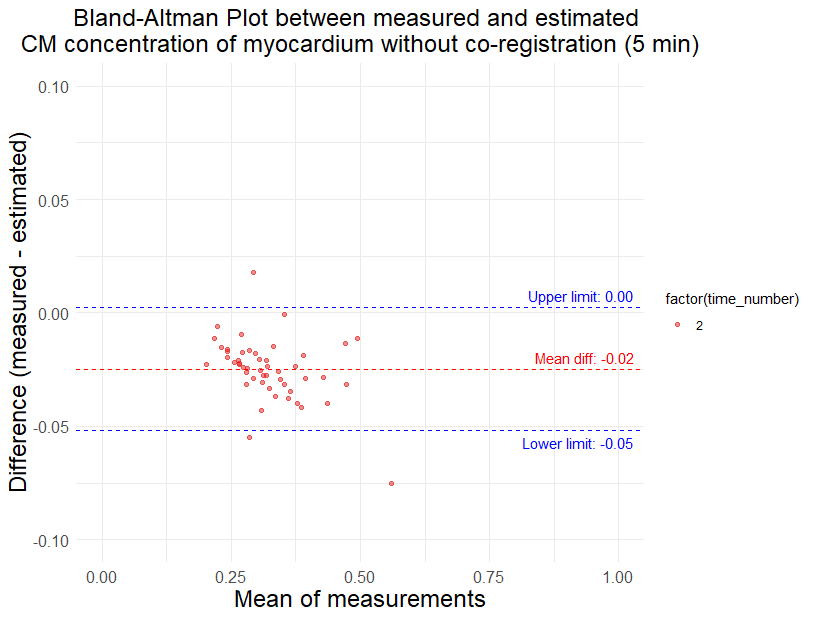

**Supplementary Fig. S4**

Bland-Altman plot comparing measured and estimated contrast medium concentration of myocardium with (upper) or without (bottom) image co-registration, 9 minutes after injection. *CM* contrast medium

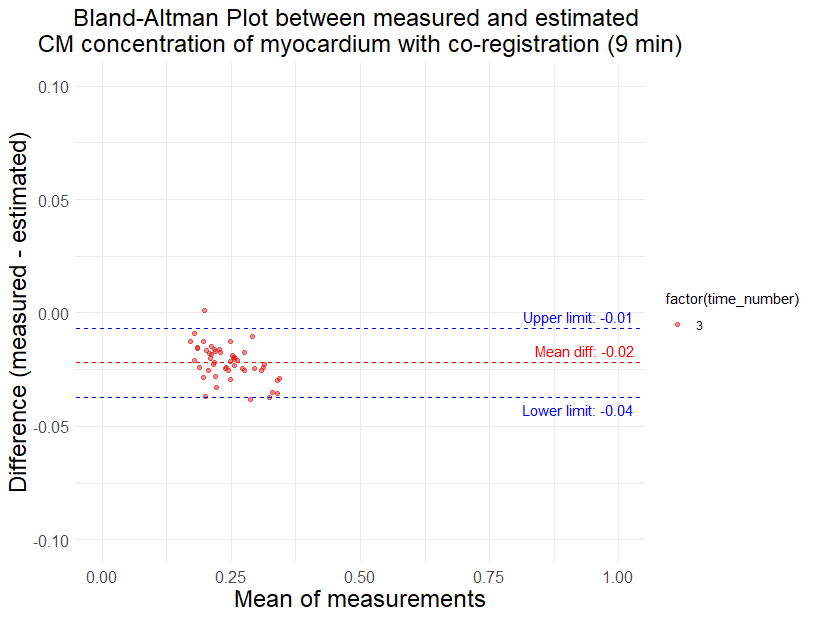

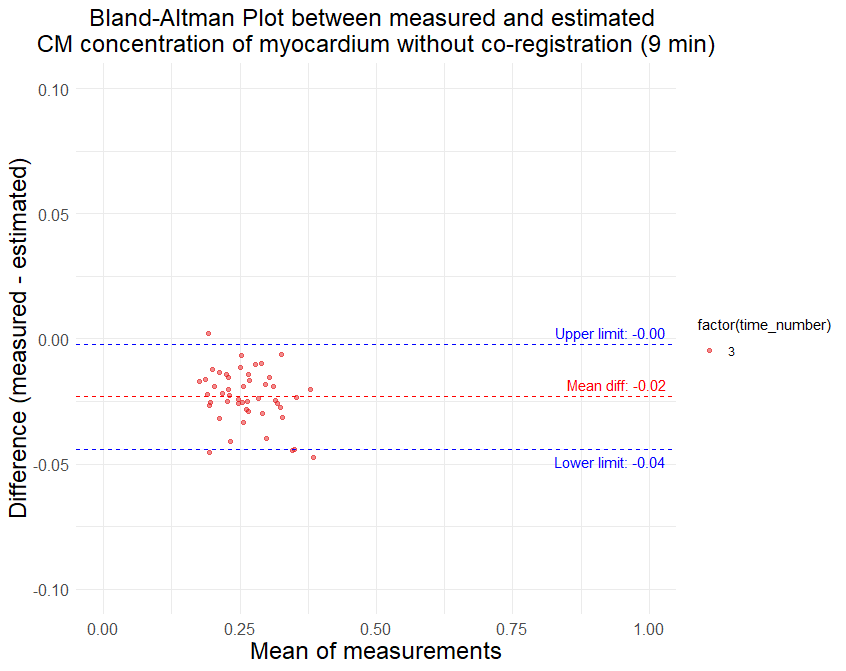

**Supplementary Fig. S5**

Bland-Altman plot comparing measured and estimated contrast medium concentration of myocardium with (upper) or without (bottom) image co-registration, 15 minutes after injection. *CM* contrast medium

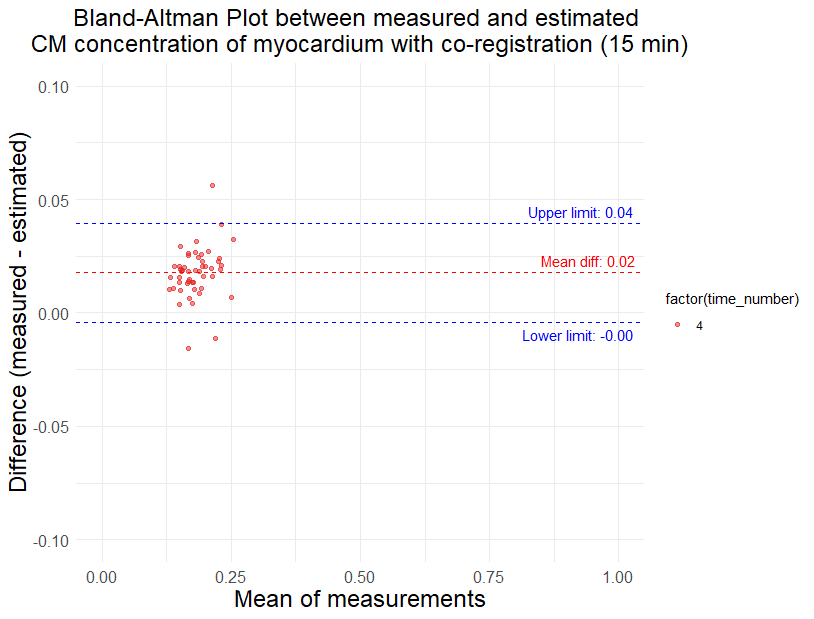

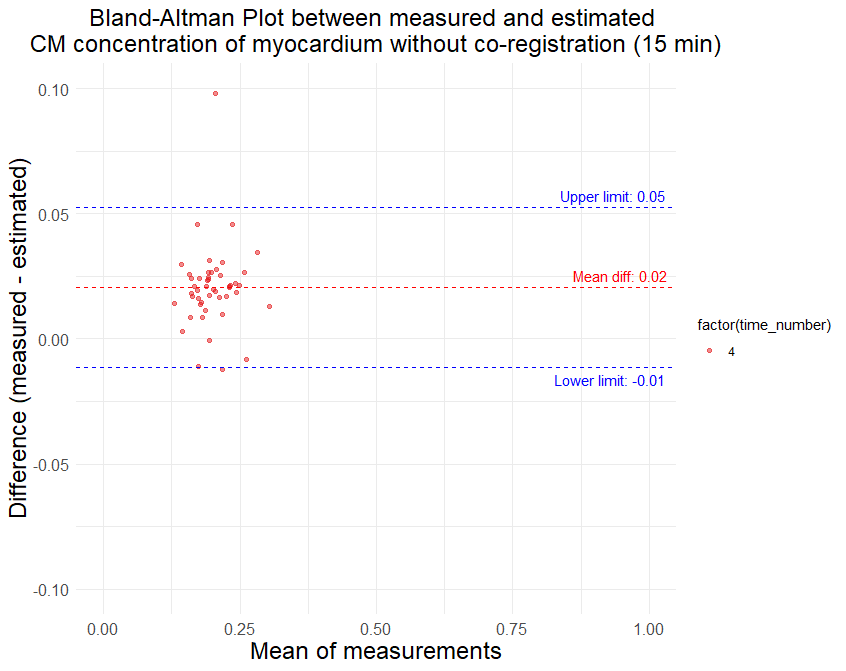

**Supplementary Fig. S6**

Bland-Altman plot comparing measured and estimated contrast medium concentration of left ventricular blood pool with (upper) or without (bottom) image co-registration, 2 minutes after injection. *CM* contrast medium, *LV* left ventricle

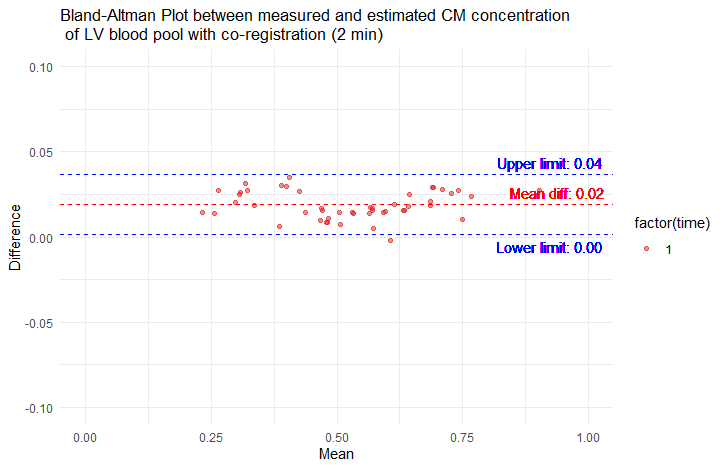

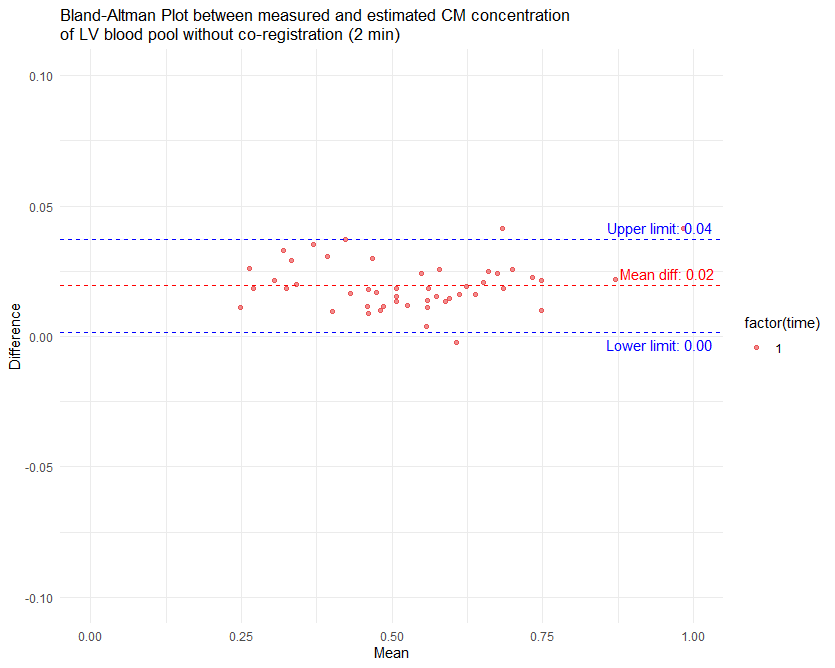

**Supplementary Fig. S7**

Bland-Altman plot comparing measured and estimated contrast medium concentration of left ventricular blood pool with (upper) or without (bottom) image co-registration, 5 minutes after injection. *CM* contrast medium, *LV* left ventricle

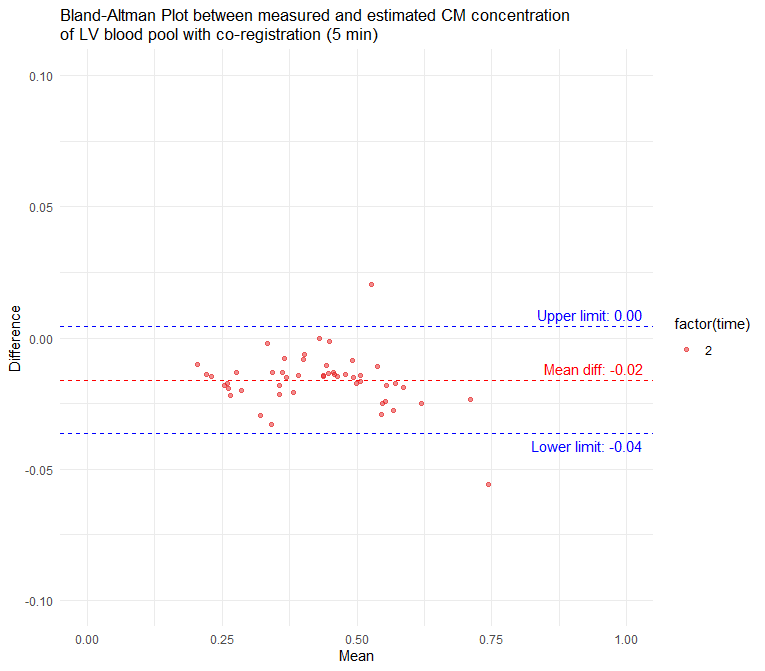

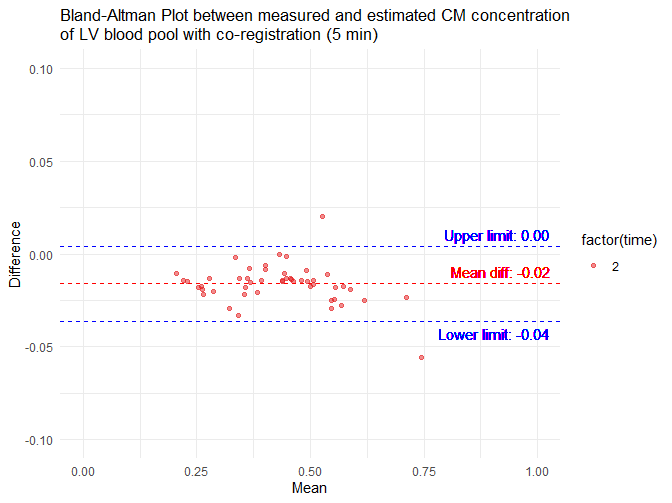

**Supplementary Fig. S8**

Bland-Altman plot comparing measured and estimated contrast medium concentration of left ventricular blood pool with (upper) or without (bottom) image co-registration, 9 minutes after injection. *CM* contrast medium, *LV* left ventricle

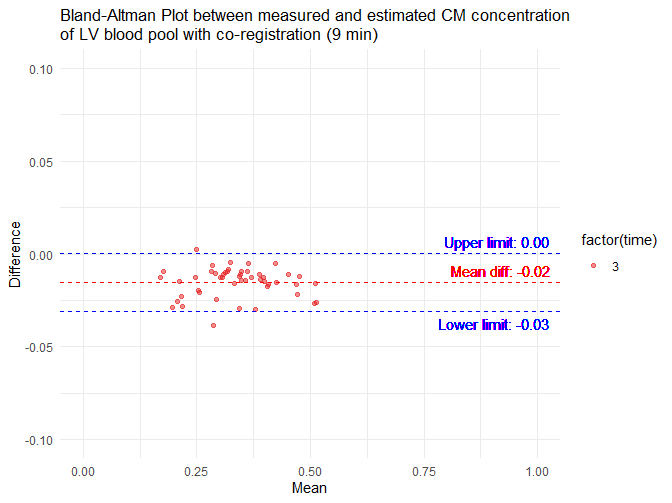

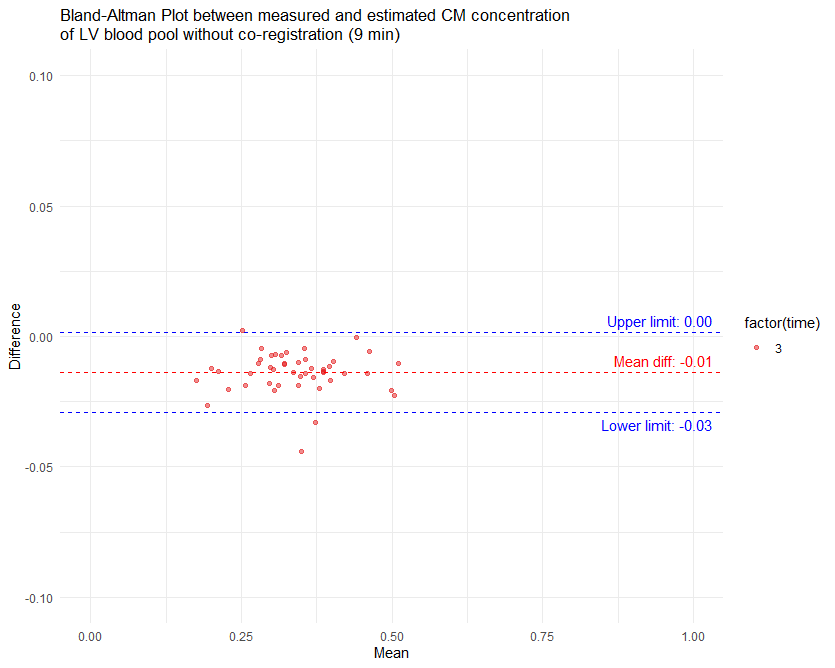

**Supplementary Fig. S9**

Bland-Altman plot comparing measured and estimated contrast medium concentration of left ventricular blood pool with (upper) or without (bottom) image co-registration, 15 minutes after injection. *CM* contrast medium, *LV* left ventricle

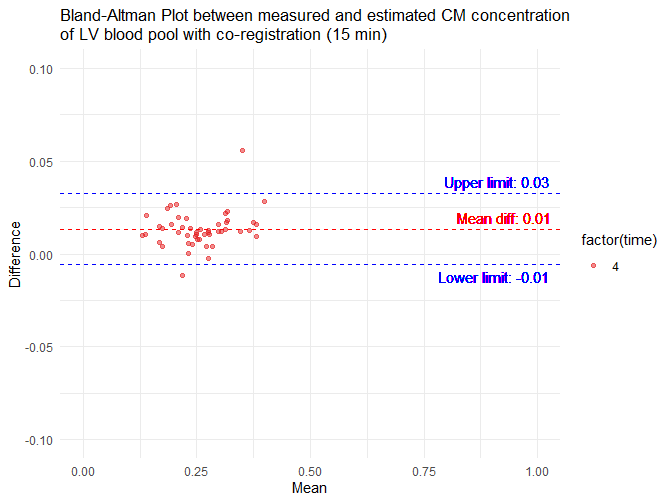

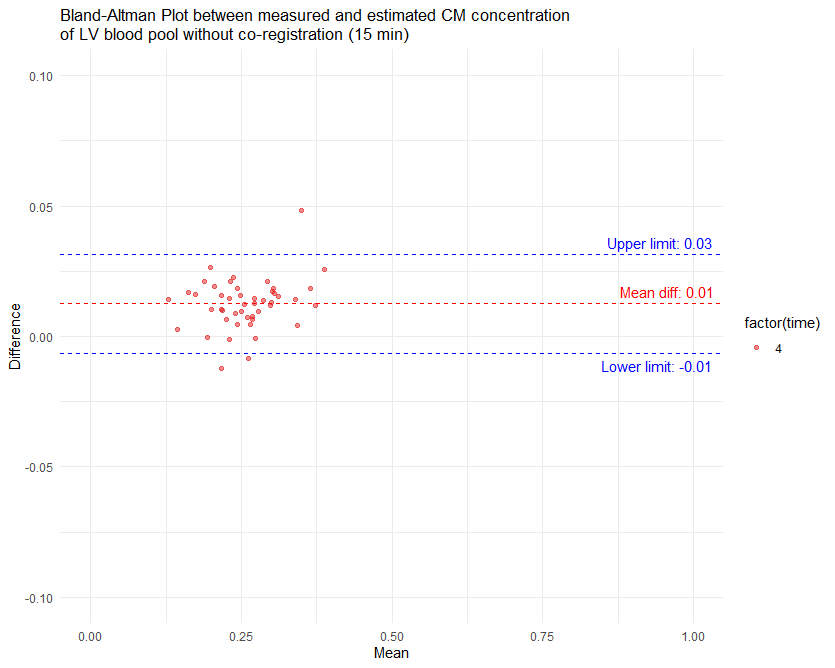
